# Predictive Value of V̇O_2peak_ in Adult Congenital Heart Disease in Comparison with Heart Failure with Reduced Ejection Fraction

**DOI:** 10.1101/2024.10.11.24315308

**Authors:** Andrea Soares, Lauren K. Park, Emily Mansour, Elena Deych, Alyssa Puritz, Min Zhao, Chao Cao, Andrew R. Coggan, Phillip M Barger, Randi Foraker, Susan B Racette, Linda R Peterson

**Affiliations:** Department of Medicine; Cardiovascular Division, Washington University School of Medicine, St. Louis, MO; Department of Internal Medicine - Pediatrics, University of Texas Health Science Center at Houston, Houston, TX; Division of General Medical Sciences, Washington University School of Medicine, St. Louis, MO; Department of Kinesiology, Indiana University Indianapolis, Indianapolis, IN; Indiana Center for Musculoskeletal Health, Indiana University School of Medicine, Indianapolis, IN; College of Health Solutions, Arizona State University, Phoenix, AZ

**Author notes:** Address for correspondence: Linda R. Peterson, MD Washington University School of Medicine MSC 8086-043-13 660 S. Euclid Ave. St. Louis, MO 63110 Facsimile.

## Abstract

**BACKGROUND:** Peak oxygen consumption (V̇O_2peak_) is used to predict outcomes and the timing of transplantation in patients with heart failure with reduced ejection fraction (HFrEF); V̇O_2peak_ also has predictive utility in patients with adult congenital heart disease (ACHD). However, the predictive value of a given V̇O_2peak_ in patients with ACHD compared to those with HFrEF, especially after adjustment for age and sex, is not clear.

**METHODS:** To address this, we performed a longitudinal cohort study comparing patients with ACHD to patients with HFrEF. The ACHD and HFrEF cohorts were matched for sex and age (+/- 10 y). V̇O_2peak_ tests were conducted between 1993 and 2012. Events were defined as death, cardiac transplantation, or left ventricular assist device placement. Outcome data were obtained via electronic medical record, Social Security Death Index, and phone interview. Cox proportional-hazard regressions were used to evaluate relationships of event-free survival with predictor variables.

**RESULTS:** Patients with ACHD (N=137) and HFrEF (N=137) with a median follow-up time of 14.5 (13.4-15.6) y in the ACHD cohort and 19 (14.8-21.1) y in the HFrEF cohort. Higher V̇O_2peak_ was associated with lower risk for a cardiac outcome, independent of age and sex, in both ACHD (HR 0.89, 95% CI 0.83-0.96, *P*=0.002) and HFrEF (HR 0.85, 95% CI 0.81-0.89, *P* <0.001Male sex was associated with greater risk of a cardiac outcome (*P*=0.001) in ACHD (HR 3.34) and HFrEF (HR 1.83). After multivariable adjustment (that included age, sex, and V̇O_2peak_) having ACHD conferred a 66% lower risk of a cardiovascular event compared to a HFrEF diagnosis (HR 0.34, 95% CI 0.22-0.53, *P*<0.001).

**CONCLUSIONS:** V̇O_2peak_ independently predicts event-free survival among adults with ACHD or HFrEF and has clinical utility in the outpatient setting. Patients with ACHD, however, have a better prognosis for any given V̇O_2peak_ compared to those with HFrEF.

**WHAT IS NEW?:**

- In an age- and sex-matched longitudinal cohort study with over 7 y of follow-up, adults with congenital heart disease (ACHD) were found to have a better event-free (no transplant or LVAD) survival than adults with heart failure with reduced ejection fraction (HFrEF) even after multivariable adjustment that included age, sex, and V̇O_2peak_. Thus, for any given V̇O_2peak_ a better event-free survival would be expected in ACHD compared with HFrEF. For both groups, a higher V̇O_2peak_ did still confer an improved event-free survival and male sex conferred a worse event-free survival.

**WHAT ARE THE CLINICAL IMPLICATIONS?:** Patients with HFrEF commonly undergo V̇O_2peak_ testing to evaluate clinical status, exercise capabilities, and timing for transplantation. Less commonly, patients with ACHD undergo V̇O_2peak_ testing. This study confirmed that a higher V̇O_2peak_ is still an excellent predictor of freedom from cardiac events and survival in both groups; however, for a given V̇O_2peak_, a patient with ACHD would be expected to have a markedly improved event-free survival vs. a patient with HFrEF even after adjusting for age and sex. Moreover, our analysis adds to the understanding of how much of an advantage a higher V̇O_2peak_ confers for each mL·min^-1^·kg^-1^ confers in each group, with a slightly greater incremental benefit for the ACHD group. This finding has implications for timing of referral to cardiac transplantation for patients with ACHD. Future studies are needed to determine the optimal V̇O_2peak_ cut-off for transplantation for those with ACHD. Furthermore, more studies are needed to investigate the potential mechanism(s) for the ACHD survival advantage.

---

A growing number of patients with congenital heart disease are surviving into adulthood (1–3). Patients with several types of uncorrected adult congenital heart disease (ACHD) are at increased risk of heart failure. Moreover, even successful surgical treatment does not cure all ACHD. Many patients remain at increased risk for heart failure after surgery. As the prevalence of ACHD rises, there is a corresponding increase in the number of ACHD patients who develop heart failure (1–3). In fact, among patients with repaired congenital heart disease, heart failure is the leading cause of death (4).

However, because right ventricular disease, valve dysfunction, shunting, and/or pulmonary hypertension often play a larger role in cardiac dysfunction compared to typical heart failure with left ventricular dysfunction, transplant evaluation of ACHD patients cannot be extrapolated from more established guidelines for heart failure (4). Compared to the general heart transplant population, ACHD patients tend to be younger, leaner, and have fewer medical comorbidities (3). Because of pulmonary dysfunction, these patients often require heart-lung transplantation. The transplant outcomes are also different among patients with ACHD: mortality 30 d after cardiac transplant is higher but long-term survival is better than in the general heart transplant population (3).

Cardiopulmonary exercise testing (CPET) provides detailed information about whole body oxygen metabolism and functional capacity through measurement of respiratory gas exchange, which is especially useful for predicting prognosis in patients with heart failure (5–10). Peak oxygen consumption (V̇O_2peak_) during maximal exercise is a standard measure used clinically to improve timing of transplantation and to objectively assess physical capacity. In patients with heart failure with reduced ejection fraction (HFrEF) who are not taking beta-blockers, a V̇O_2peak_ value <14 mL·min^-1^·kg^-1^ (<12 mL·min^-1^·kg^-1^ in those taking beta-blockers) is one criterion used to indicate that a patient would benefit from a heart transplant (5, 6). Although V̇O_2peak_ has been demonstrated to have excellent prognostic value among patients with HFrEF, relatively little is known regarding the prognostic value of CPET among patients with ACHD (11). The European Association for Cardiovascular Prevention & Rehabilitation and the American Heart Association recommend that CPET be used among patients with ACHD (12). However, the implications of a given V̇O_2peak_ are amongst patients with ACHD are unclear, especially in relation to the more well-known implications of a particular V̇O_2peak_ in patients with HFrEF (5,6).

Thus, the first aim of this longitudinal study was to investigate the prognostic value of V̇O_2peak_ among patients with ACHD and a sex- and age-matched (within ten years) cohort of patients with non-congenital HFrEF. A second aim was to compare the cardiac event implications between patients with ACHD and those with HFrEF after adjustment for V̇O_2peak_. Our hypotheses were that higher V̇O_2peak_ would be predictive of improved event-free survival in both cohorts, and that the patients with ACHD would have longer event-free survival compared to patients with HFrEF even after adjustment for V̇O_2peak_.

## METHODS

### Study Design and Population

This was a longitudinal, observational, case-matched study. Data were reviewed from 1251 patients who were referred by their cardiologist for V̇O_2peak_ testing and completed the test in our laboratory at Washington University School of Medicine between May 1993 and December 2012. All patients had a minimum of 7 y of follow up for end point determination. Clinical data and medications were recorded on the day of the V̇O_2peak_ test. This study was approved by the Washington University in St. Louis Institutional Review Board. All patients provided written informed consent.

Eligibility criteria for inclusion in the analysis were adults with congenital heart disease (ACHD cohort) or adults with HFrEF of non-congenital origin (HFrEF cohort). ACHD diagnosis was confirmed by chart review and classified as mild, moderate, or severe according to the American Heart Association Adult Congenital Heart Disease Anatomic and Physiologic Classification (13). HFrEF was defined clinical evidence of heart failure and an ejection fraction <45%, determined by echocardiography within one year of the V̇O_2peak_ test.

### Cardiopulmonary Exercise Testing

All patients underwent a symptom-limited incremental exercise test on a treadmill. Height and weight were measured before the test utilizing a stadiometer and digital scale. The starting treadmill speed was based on the patient’s fitness level and the grade was 0%. Increases in treadmill speed and grade during the test were individualized to promote reaching V̇O_2peak_ in ∼8-12 min. Standard open-circuit spirometry was performed to measure respiratory gas exchange using a calibrated metabolic cart (ParvoMedics; Salt Lake City, UT). Fractional concentrations of expired oxygen and carbon dioxide were quantified from a mixing chamber using electronic gas analyzers. Oxygen consumption (V̇O_2_), carbon dioxide production (V̇CO_2_), respiratory exchange ratio (V̇CO_2_/V̇O_2_), ventilation, heart rate, blood pressure, and rating of perceived exertion (RPE) were obtained throughout the test, as described previously (6). V̇O_2peak_ was determined by averaging the highest consecutive V̇O_2_ values over a 1 min period at the end of the exercise test. Ventilatory equivalent was calculated as the ratio of ventilation (L/min) to V̇O_2peak_ (L/min) at peak exercise. Oxygen pulse (mL/beat) was calculated as the ratio of V̇O_2peak_ (mL/min) to heart rate (bpm) at peak exercise, which reflects the oxygen consumption in all body tissues per each heartbeat.

### Cardiac Events

The primary end point was a cardiac event, defined as patient death, heart transplant, or left ventricular assist device (LVAD) placement. Information about actuarial status and dates of death, heart transplant, and LVAD placement were obtained via electronic medical record, Social Security Death Index, or phone interview. A cardiac event was considered a censoring event.

### Statistical Analysis

To investigate the association between the prognostic implication of V̇O_2peak_ values and event-free survival in patients with ACHD relative to those with HFrEF, a retrospective, matched comparison study was performed. Patients in the congenital cohort and HFrEF cohort were sex-matched and age-matched within 10 y. Data are shown as median [interquartile range], mean ± standard deviation, or proportion of the sample. Baseline characteristics were compared between cohorts using Student’s unpaired t-tests for continuous variables or Wilcoxon non-parametric test, as appropriate and Chi-square tests for categorical variables. The groups were compared visually and statistically using Kaplan-Meier curves with log-rank tests. Cox-proportional hazard regression models were then used to investigate the association between survival time for each cohort of patients and one or more predictor variables. Event-free survival was defined as survival free of a heart transplant or LVAD placement. Proportionality assumptions were tested for Cox models All analyses were performed using R statistical software version 4.3.2 A *P* value <0.05 was considered statistically significant.

## RESULTS

### Patient Population

After sex- and age-matching of the HFrEF patients to the ACHD patients, there were 137 patients in the ACHD cohort and an equal number in the HFrEF cohort for analysis. Of the HFrEF patients the vast majority had nonischemic cardiomyopathy (Table 1). Censoring-adjusted median follow-up time was 19.0 (14.8-21.1) y in the ACHD cohort and 14.5 (13.4-15.6) y in the HFrEF cohort Baseline characteristics of both cohorts are shown in Table 1. In the ACHD cohort, 92% had moderate or severe disease. Despite performing age-matching within 10 y, age was lower in the ACHD cohort, as was body mass index. A higher proportion of the HFrEF cohort was taking angiotensin-converting enzyme inhibitors, angiotensin-receptor blockers, and β-blockers.

**Table 1.** Baseline Characteristics

|  | **ACHD** | **HFrEF** | *P* value |
| --- | --- | --- | --- |
| n | 137 | 137 |  |
| Age at $\dot{V}O_{2peak}$ test (years) | 30.8 [25.2,39.8] | 40.3 [32.7,46.0] | **< 0.001** |
| Sex: male / female (%) | 55% / 45% | 55% / 45% | NS |
| Race: white / black (%) | 90% / 10% | 70% / 30% | **< 0.001** |
| Body Mass Index (kg/m<sup>2</sup>) | 24.6 [21.9,28.2] | 28.8 [24.7,33.2] | **< 0.001** |
| Medications (% taking) |  |  |  |
| ACE Inhibitor | 40% | 61% | **< 0.001** |
| ARB | 7% | 19% | **0.004** |
| β-Blocker | 26% | 75% | **< 0.001** |
| ACHD disease complexity (%) |  |  |  |
| Simple | 8% |  |  |
| Moderate | 52% |  |  |
| Complex | 40% |  |  |
| Ischemic heart failure (%) | 0% | 23% | NS |

Data are expressed as median [interquartile range] or as a proportion of the sample. Bold *p* values reflect statistical significance. ACE, angiotensin-converting enzyme; ACHD, adult congenital heart disease; ARB, angiotensin II receptor blocker; HFrEF, heart failure with reduced ejection fraction.
**Student's unpaired t-test** was used to evaluate between-group differences of continuous variables. **Chi-square tests** were used to analyze between-group differences for categorical variables.

As shown in Table 2, V̇O_2peak_ was significantly higher in the ACHD cohort compared to the HFrEF cohort, whether expressed relative to body mass or in absolute terms. Nearly identical median RER peak values indicate that in general, patients in both groups gave maximal or near-maximal effort. Peak ventilation, oxygen pulse, maximum heart rate and maximum systolic and diastolic blood pressures were significantly higher in the ACHD cohort.

**Table 2.**
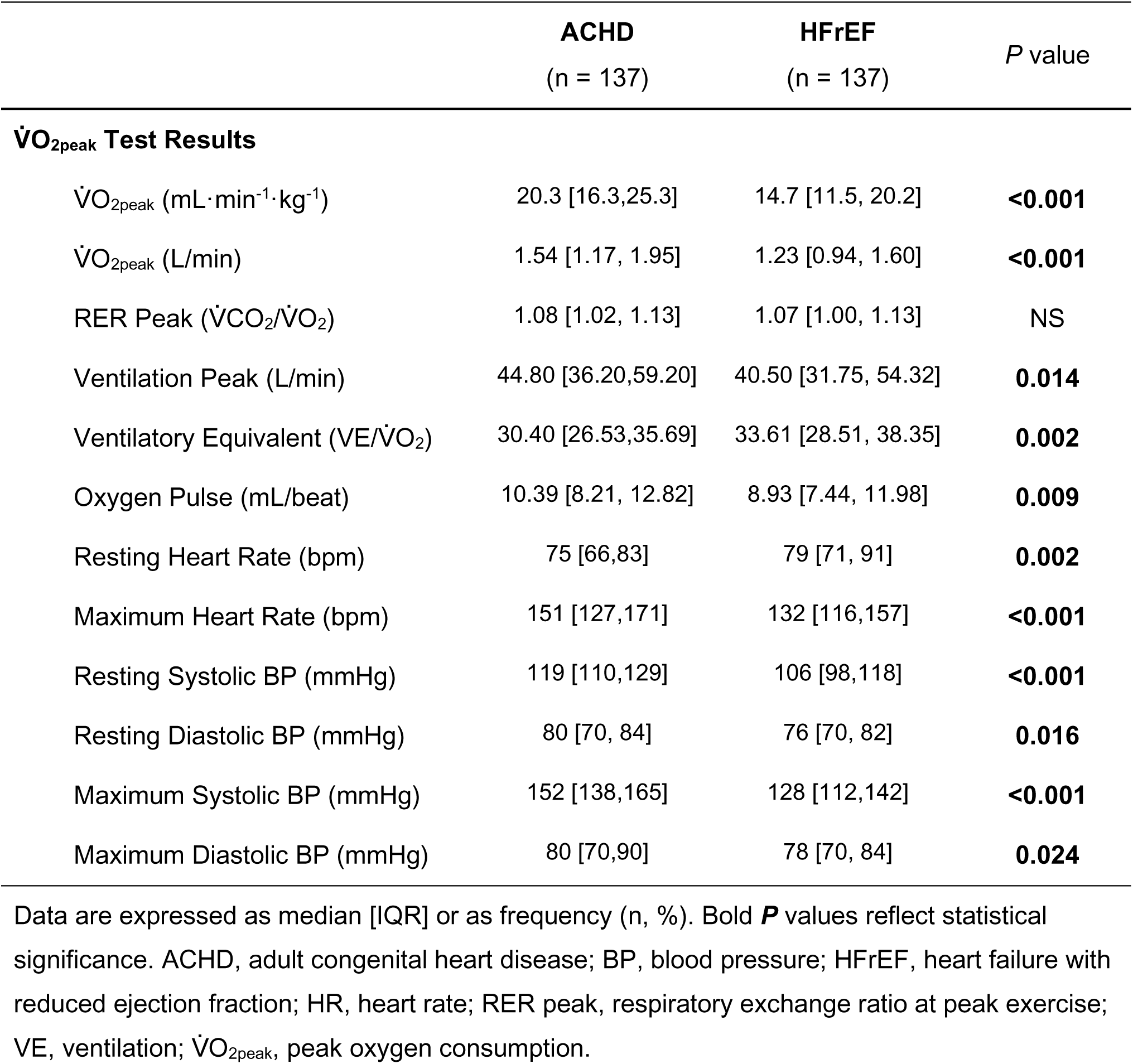
V̇O_2peak_ Test Results and Outcome Frequency

|  | **ACHD**<br>(n = 137) | **HFrEF**<br>(n = 137) | *P* value |
| --- | --- | --- | --- |
| **$\dot{V}O_{2peak}$ Test Results** |  |  |  |
| $\dot{V}O_{2peak}$ (mL·min<sup>-1</sup>·kg<sup>-1</sup>) | 20.3 [16.3,25.3] | 14.7 [11.5, 20.2] | **<0.001** |
| $\dot{V}O_{2peak}$ (L/min) | 1.54 [1.17, 1.95] | 1.23 [0.94, 1.60] | **<0.001** |
| RER Peak ($\dot{V}CO_2/\dot{V}O_2$) | 1.08 [1.02, 1.13] | 1.07 [1.00, 1.13] | NS |
| Ventilation Peak (L/min) | 44.80 [36.20,59.20] | 40.50 [31.75, 54.32] | **0.014** |
| Ventilatory Equivalent (VE/$\dot{V}O_2$) | 30.40 [26.53,35.69] | 33.61 [28.51, 38.35] | **0.002** |
| Oxygen Pulse (mL/beat) | 10.39 [8.21, 12.82] | 8.93 [7.44, 11.98] | **0.009** |
| Resting Heart Rate (bpm) | 75 [66,83] | 79 [71, 91] | **0.002** |
| Maximum Heart Rate (bpm) | 151 [127,171] | 132 [116,157] | **<0.001** |
| Resting Systolic BP (mmHg) | 119 [110,129] | 106 [98,118] | **<0.001** |
| Resting Diastolic BP (mmHg) | 80 [70, 84] | 76 [70, 82] | **0.016** |
| Maximum Systolic BP (mmHg) | 152 [138,165] | 128 [112,142] | **<0.001** |
| Maximum Diastolic BP (mmHg) | 80 [70,90] | 78 [70, 84] | **0.024** |

Data are expressed as median [IQR] or as frequency (n, %). Bold ***P*** values reflect statistical significance. ACHD, adult congenital heart disease; BP, blood pressure; HFrEF, heart failure with reduced ejection fraction; HR, heart rate; RER peak, respiratory exchange ratio at peak exercise; VE, ventilation; $\dot{V}O_{2peak}$ , peak oxygen consumption.

The most common cardiac event for both groups was death (Table 2), with more than double the number of deaths in the HFrEF group as compared with the ACHD group. There were very few heart transplants or LVAD placements in the ACHD group overall; thus, there were >8 times as many HFrEF patients who had undergone a heart transplant and 11 times as many HFrEF patients who underwent LVAD placement in comparison with the ACHD group.

Results of the multivariable Cox regression analysis for all patients and for each cohort are shown in Table 3. Overall, an ACHD diagnosis was associated with marked protection against a cardiac event as compared with a HFrEF diagnosis, independent of age, sex, and V̇O_2peak_ (HR 0.29, *P*< 0.001). A higher V̇O_2peak_ was associated with lower risk for a cardiac outcome, independent of age and sex in both cohorts together and in each cohort separately. For the ACHD cohort each increase of 1 mL·min^-1^·kg^-1^ was associated with a 13% (CI 6.0%-19.9%, *P*<0.001;) better event-free survival, and for the HFrEF cohort, each incremental increase in V̇O_2peak_ was associated with a 15% (CI 11%-19%, *P*=0.001). Male sex was associated with doubling of risk of a cardiac event when both cohorts were combined and in each cohort . Overall, in both groups there was also a cardiac event-free survival advantage in those taking beta-adrenergic blocking agents.

**Table 3.**
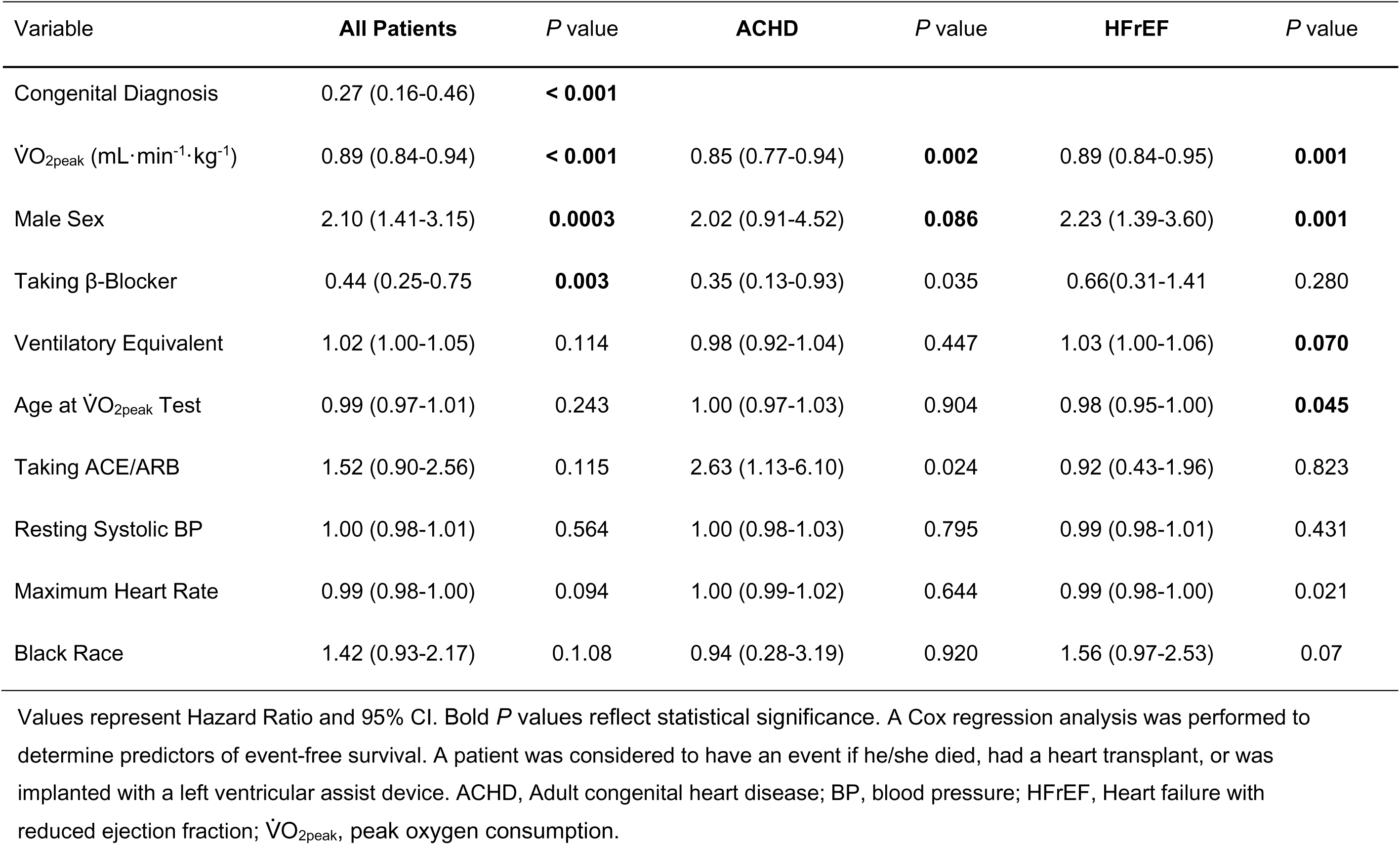
Multivariable Cox regression analysis of risk factors for a cardiac event

| Variable | All Patients | *P* value | ACHD | *P* value | HFrEF | *P* value |
| --- | --- | --- | --- | --- | --- | --- |
| Congenital Diagnosis | 0.27 (0.16-0.46) | **< 0.001** |  |  |  |  |
| $\dot{V}O_{2peak}$ (mL·min<sup>-1</sup>·kg<sup>-1</sup>) | 0.89 (0.84-0.94) | **< 0.001** | 0.85 (0.77-0.94) | **0.002** | 0.89 (0.84-0.95) | **0.001** |
| Male Sex | 2.10 (1.41-3.15) | **0.0003** | 2.02 (0.91-4.52) | **0.086** | 2.23 (1.39-3.60) | **0.001** |
| Taking β-Blocker | 0.44 (0.25-0.75) | **0.003** | 0.35 (0.13-0.93) | 0.035 | 0.66(0.31-1.41) | 0.280 |
| Ventilatory Equivalent | 1.02 (1.00-1.05) | 0.114 | 0.98 (0.92-1.04) | 0.447 | 1.03 (1.00-1.06) | **0.070** |
| Age at $\dot{V}O_{2peak}$ Test | 0.99 (0.97-1.01) | 0.243 | 1.00 (0.97-1.03) | 0.904 | 0.98 (0.95-1.00) | **0.045** |
| Taking ACE/ARB | 1.52 (0.90-2.56) | 0.115 | 2.63 (1.13-6.10) | 0.024 | 0.92 (0.43-1.96) | 0.823 |
| Resting Systolic BP | 1.00 (0.98-1.01) | 0.564 | 1.00 (0.98-1.03) | 0.795 | 0.99 (0.98-1.01) | 0.431 |
| Maximum Heart Rate | 0.99 (0.98-1.00) | 0.094 | 1.00 (0.99-1.02) | 0.644 | 0.99 (0.98-1.00) | 0.021 |
| Black Race | 1.42 (0.93-2.17) | 0.1.08 | 0.94 (0.28-3.19) | 0.920 | 1.56 (0.97-2.53) | 0.07 |

Values represent Hazard Ratio and 95% CI. Bold *P* values reflect statistical significance. A Cox regression analysis was performed to determine predictors of event-free survival. A patient was considered to have an event if he/she died, had a heart transplant, or was implanted with a left ventricular assist device. ACHD, Adult congenital heart disease; BP, blood pressure; HFrEF, Heart failure with reduced ejection fraction; $\dot{V}O_{2peak}$ , peak oxygen consumption.

The Kaplan-Meier plot depicting event-free survival for all patients as a function of V̇O_2peak_ is shown in Figure 2. A V̇O_2peak_ cut point of 14 mL·min^-1^·kg^-1^ was clinically meaningful, with values >14 mL·min^-1^·kg^-1^ reflecting longer event-free survival compared to V̇O_2peak_ values ≤14. The Kaplan-Meier plot in Figure 3 shows survival curves by cohort and V̇O_2peak_; these curves depict cardiac event-free survival was longer in the ACHD cohort than in the HFrEF cohort, similar to what was seen in the multivariate analysis in Table 3. Furthermore, the V̇O_2peak_ cut point of 14 mL·min^-1^·kg^-1^ remained statistically significant? in both cohorts, with values >14 reflecting longer event-free survival than values ≤14 mL·min^-1^·kg^-1^ among ACHD and HFrEF patients alike.

**Figure 1.**
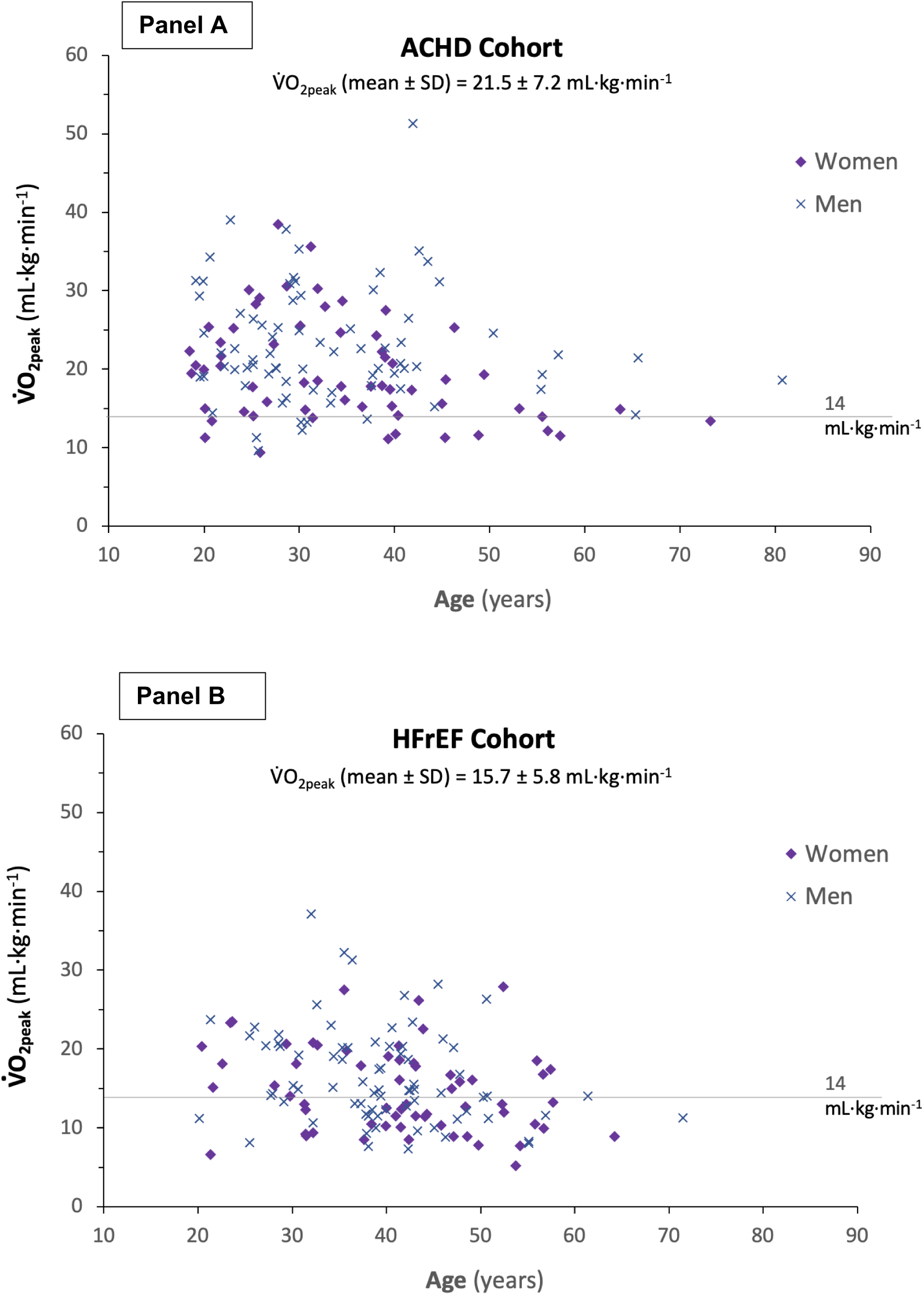
Baseline V̇O_2peak_ values by cohort, sex, and age. Scatterplot showing individual V̇O_2peak_ values at baseline for patients with adult congenital heart disease (ACHD, panel A) and patients with heart failure with reduced ejection fraction (HFrEF, panel B) by age (x axis). Values for women are depicted by purple diamonds and values for men are depicted by blue Xs. The horizontal line at 14 mL·min^-1^·kg^-1^ represents the threshold below which patients with HFrEF generally benefit from a heart transplant.

**Figure 2.**
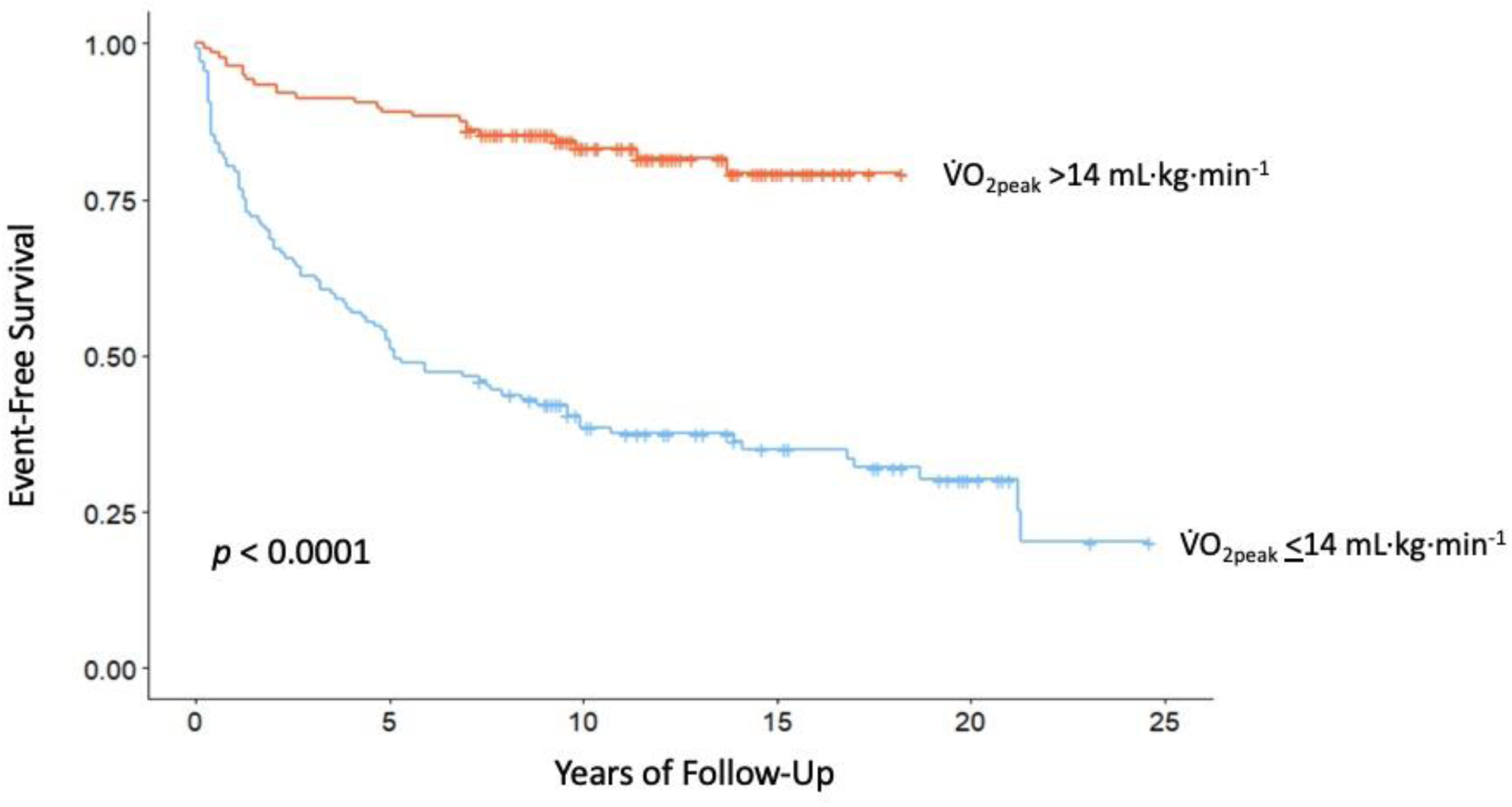
Event-free survival as a function of V̇O_2peak_ among all patients. Survival curves are shown for patients with V̇O_2peak_ <14 mL·min^-1^·kg^-1^ and patients with V̇O_2peak_ >14 mL·min^-1^·kg^-1^. Patients in the ACHD and HFrEF cohorts are combined. The difference in event-free survival between these two V̇O_2peak_ groups was significant (*P*< 0.0001).

**Figure 3.**
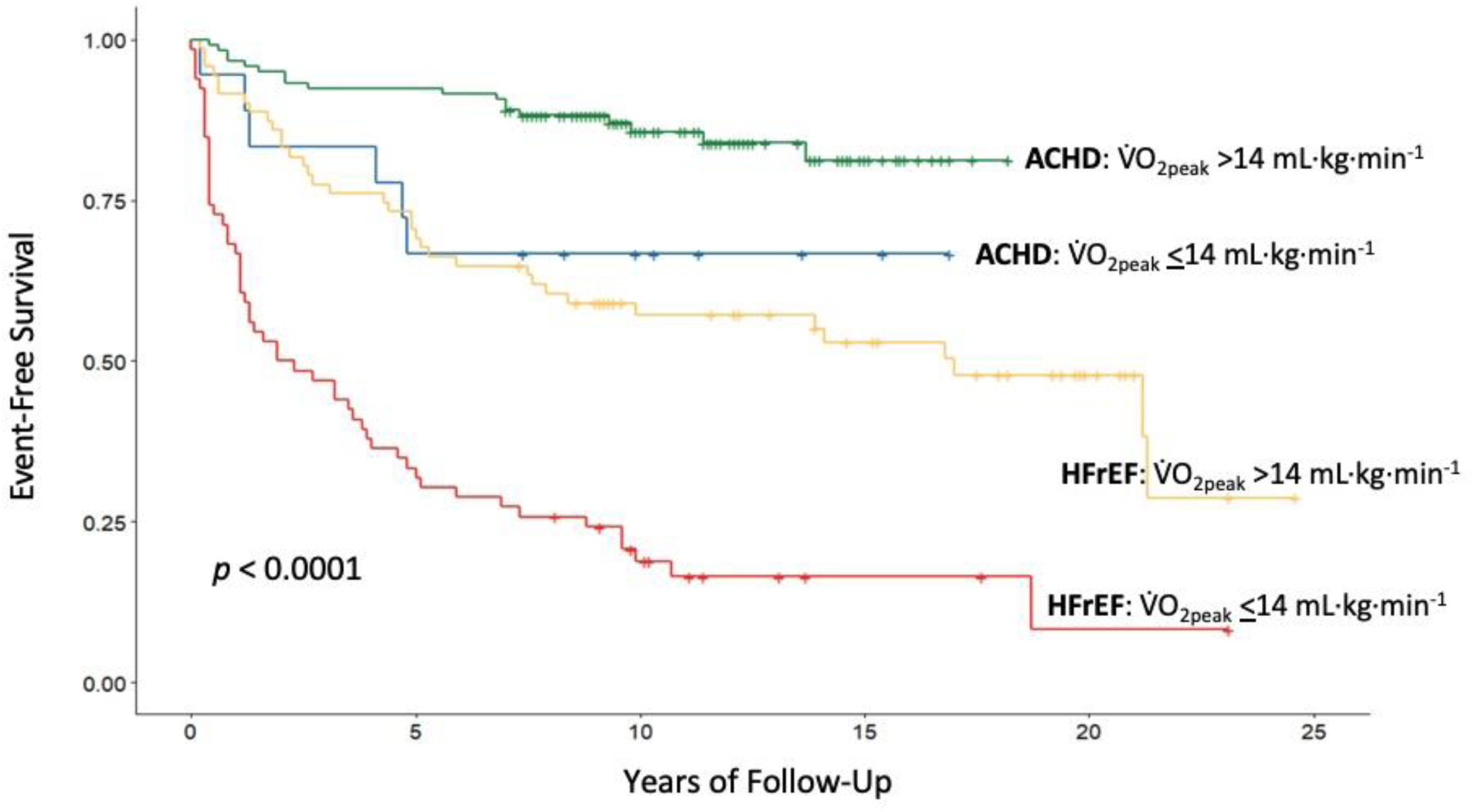
Event-free survival by cohort and V̇O_2peak_. Survival curves are shown for patients with adult congenital heart disease (ACHD) and patients with heart failure with reduced ejection fraction (HFrEF), with each cohort dichotomized by V̇O_2peak_ (< or > 14 mL·min^-1^·kg^-1^). The difference in event-free survival among the four cohorts / V̇O_2peak_ groups was significant (*P*< 0.0001).

## DISCUSSION

To our knowledge this study is the first to compare the prognostic value of V̇O_2peak_ in an ACHD cohort versus in age- and sex-matched patients with HFrEF, in whom this measurement is more commonly made for transplantation evaluation. For the ACHD patients, as well as the HFrEF patients, V̇O_2peak_ was an independent predictor of cardiac event-free survival, with an improvement for each 1 mL·min^-1^·kg^-1^ that was similar but slightly greater in the ACHD group. Most striking was the marked cardiac event-free survival advantage for those in the ACHD group vs. those with HFrEF even after matching for sex and for age-within 10 y and after accounting for these and other variables in the multivariable analyses. To wit, despite the ACHD group having a longer median follow-up time, there were ∼4 times fewer cardiac events and half the number of deaths in the ACHD group than in the HFrEF group. In both cohorts, male sex was associated with greater risk for a cardiac outcome, which was more pronounced among patients with ACHD. In contrast, beta-adrenergic blocker therapy predicted a lower risk in both groups combined. In sum, the predictors of survival were V̇O_2peak_, female sex, beta-blocker use, and most dramatically, an ACHD diagnosis as opposed to HFrEF.

The results of this study support our hypothesis that not only is V̇O_2peak_ predictive of cardiac events but also that an ACHD diagnosis would confer a cardiac event-free survival advantage. This marked advantage persisted even after matching for sex and age within 10 y and multivariable adjustment. A previous study of 335 patients with ACHD and 40 patients with chronic HF demonstrated that, when matched by New York Heart Association (NYHA) class (a common metric of physical limitations in HF), ACHD patients and HF patients had similar V̇O_2peak_ values.^16^ Unlike in our study, however, these groups were not case-matched by sex or age (i.e., ACHD: 55% male; mean age 33 y; HF: 72% male; mean age 58 y) and HF was not restricted to HFrEF, which likely impacted the results. Though patients with ACHD may be less likely (than those with HFrEF) to undergo transplantation since they may need a heart-lung bloc due to Eisenmenger’s, and those with ACHD may be less likely to be offered an LVAD due to right heart failure, there is still a two-fold advantage in terms of mortality for those with ACHD over those with HFrEF, despite a longer follow-up time for the former. The reasons for this advantage are not clear. There were some baseline differences between the groups in terms of age, race, body mass index, and percentage of patients taking cardiac medications. However, age and race and beta-blocker status were included in the multivariable analyses of outcomes and V̇O_2peak_ is adjusted for weight. Moreover, several studies have shown an ‘obesity paradox’ for HFrEF in which patients with a higher BMI often have a better prognosis. Future studies are needed to determine the mechanisms explaining the marked difference in outcomes between the two groups. Regardless, given that patients with ACHD have a better event-free survival, specific guidelines for V̇O_2peak_-guided treatment in the ACHD population are warranted.

The results of this study also support our hypothesis that higher V̇O_2peak_ would be predictive of improved event-free survival in both cohorts. The observation that V̇O_2peak_ is an independent predictor of outcomes in HFrEF patients is consistent with previous studies both in the pre- and post-beta-blocker eras.^5,6^ The data showing that V̇O_2peak_ is predictive of outcomes in ACHD patients is consistent with the results of a recent systematic review and meta-analysis of 32 studies of patients with ACHD,^14^ which demonstrated the prognostic value of several peak and submaximal measures derived from a cardiopulmonary exercise test (CPET) measures for predicting major adverse cardiovascular events (MACE). There was also a recent large study of ACHD patients by Inuzuka et al. that showed the predictive utility of V̇O_2peak_.^15^ However, the follow-up period for both the meta-analysis (median 47 mo)^14^ and for the study by Inuzuka^15^ (5 y) was shorter than our follow-up period of 10 y, which is especially important given the relatively young age of patients in the ACHD studies. Moreover, our analysis adds to the understanding of how much of an advantage a higher V̇O_2peak_ confers for each 1 mL·min^-1^·kg^-1^ confers in each group, with a slightly greater incremental benefit for the ACHD group.

Another striking finding from our investigation is that male sex was associated with more than twice the risk of a cardiac outcome than female sex in patients with ACHD. Although in general women have been shown to have higher cardiovascular disease mortality compared to men, our results were consistent with prior studies in patients with ACHD indicating that male sex is associated with higher cardiac mortality in this population.^19^ Interestingly, male sex was also strongly associated with an increased risk of a cardiac event in the HFrEF cohort as was seen in our previous longitudinal study of HFrEF patients and in other studies of sex-related outcomes in these patients.

Limitations of the study include the relatively small sample size, the small proportion of non-white patients, and the restriction of the samples to a single-center. Additionally, the ACHD diagnoses were variable, although most conditions were categorized as moderate to complex according to the American Heart Association Adult Congenital Heart Disease Anatomic and Physiologic Classification.

The clinical implications of the current study for the care of patients with ACHD, as well as HFrEF, are manifold. Due to early diagnosis and advancement in surgical techniques, patients with ACHD are living longer than ever before. Thus, it is essential to continue to develop and evaluate tools to risk-stratify patients with ACHD based on objective measures of functional status. The clinical value of V̇O_2peak_ in predicting cardiac events including death in patients with ACHD highlights the utility of CPET as an important clinical tool. Our results further highlight how much of an event-free survival advantage can be expected from each 1 mL·min^-1^·kg^-1^ for patients with ACHD or HFrEF. The current study also revealed that ACHD status confers a marked independent event-free survival advantage as compared with HFrEF status. Thus, for any given V̇O_2peak_, a patient with ACHD would be expected to have a significantly better event-free survival rate as compared with a patient with HFrEF. More research is needed to evaluate the potential mechanisms underpinning the marked survival advantage that can be expected in ACHD vs. HFrEF from our findings, in addition to the known sex-related differences in survival.

## Data Availability

All data are available.

## Acknowledgments

We thank the patients for participating in this study and the staff of the Barnes-Jewish Hospital Cardiac Diagnostic Laboratory for their contributions in conducting the V̇O_2peak_ tests.

## Sources of Funding

Supported by the Mentors in Medicine Award, St. Louis, Missouri (AS), the Clinical and Translational Science Award (CTSA) Grant [UL1 TR000448], Siteman Comprehensive Cancer Center and NCI Cancer Center Support Grant P30 CA091842, and R33HL155858 (LRP, SBR, LKP, ARC), R01AG071717-01A1 (SBR, LRP), R01 AG060499-01, R01 HL165238-01A1 and AHA Second Century Early Implementation Science Award #23SCISA1145192 (LRP) and 5T32HL130357-05 (LKP).

## Disclosures

The authors have no financial disclosures that are relevant to this manuscript.

## Notes

### Competing Interest Statement

The authors have declared no competing interest.

### Author Declarations

Washington University School of Medicine IRB HRPO board provided oversight for this project.

